# Chronic fatigue syndrome, depression, and anxiety symptoms due to relapsing-remitting multiple sclerosis are associated with reactivation of Epstein-Barr virus and Human Herpesvirus 6

**DOI:** 10.1101/2024.10.12.24315393

**Authors:** Michael Maes, Abbas F. Almulla, Elroy Vojdani, Elizabet Dzhambazova, Drozdstoj Stoyanov, Yingqian Zhang, Aristo Vojdani

## Abstract

Relapsing-remitting multiple sclerosis (RRMS) is defined by elevated IgG/IgA/IgM responses targeting Epstein-Barr Virus (EBV) nuclear antigen 1 (EBNA) and deoxyuridine-triphosphatases (dUTPases) of Human herpsesvirus-6 (HHV-6) and EBV. These responses suggest that the viruses are being replicated and reactivated. An increased prevalence of chronic fatigue syndrome, depression, and anxiety is associated with signs of immune activation in RRMS. Nevertheless, there is a lack of data regarding the association between viral reactivation and neuropsychiatric symptoms of RRMS. This study investigated the IgG/IgA/IgM responses to EBNA, and EBV and HHV-6-dUTPases, in 58 remitted RRMS patients and 63 normal controls. The McDonald criteria were employed to establish the diagnosis of MS. The Expanded Disability Status Scale (EDSS) and the Multiple Sclerosis Severity Score were employed to evaluate disabilities caused by RRMS. We evaluated the scores of the Hamilton Depression (HAMD) and Anxiety (HAMA) Rating Scales, and Fibro-Fatigue (FF) scale. One latent construct was extracted from the EDSS, MSSS, FF, HAMD, and HAMA scores. We discovered that the combined effects of IgG and IgM-HHV-6-dUTPAses accounted for 63.7% of the variance in this construct. Furthermore, the total FF, HAMA, and HAMD scores were substantially associated with the IgG and IgM-HHV-6-dUTPAses, accounting for approximately 38.7% to 51.0% of the variance. The three neuropsychiatric rating scale scores were also significantly correlated with IgA reactivity directed to both dUTPases and IgG/IgA/IgM to EBNA. In conclusion, the reactivation and replication of HHV-6 and EBV significantly contributes to chronic fatigue syndrome, as well as symptoms of depression and anxiety due to RRMS.

## Introduction

Multiple sclerosis (MS) is an autoimmune disorder distinguished by a range of pathological processes, including neuroinflammation, oxidative stress-induced damage, neurodegeneration, as well as axonal and neuronal injury. Additionally, it involves excitotoxicity, demyelination, and the formation of glial scars within the nervous system (Abbasi et al., 2016; de Carvalho Jennings Pereira et al., 2020; Debouverie, 2009; Gonsette, 2008; Kallaur et al., 2016; Morris and Maes, 2013a; Papiri et al., 2023; Siotto et al., 2019; Sospedra and Martin, 2016). Multiple sclerosis manifests in a diverse array of disabilities, encompassing compromised motor function and sensory irregularities that may exhibit patterns of exacerbation and remission, particularly in cases of relapsing remitting MS (RRMS) (Almulla et al., 2023a; Jongen et al., 2012; Staff et al., 2009).

Latent viral infections, specifically those caused by Epstein-Barr virus (EBV) and human herpesvirus 6 (HHV-6), have been identified within the central nervous system (CNS) and cerebrospinal fluid of patients diagnosed with MS. The presence of these viruses correlates with disease activity, indicating their potential involvement in the pathogenesis and progression of multiple sclerosis (Engdahl et al., 2019; Khalesi et al., 2023; Lundström and Gustafsson, 2022). The levels of immunoglobulin G (IgG) that target the viral proteins of HHV-6 and EBV, particularly the EBV nuclear antigen 1 (EBNA), exhibit a correlation with the disabilities and progression observed in MS (Comabella et al., 2010; Farrell et al., 2009; Sundström et al., 2004; Villoslada et al., 2003). Elevated IgM reactivity directed towards HHV-6 (IgM-HHV-6) serves as another biomarker for MS (Villoslada et al., 2003). The heightened responses of IgG/IgA/IgM targeting the deoxyuridine-triphosphatase (dUTPase) of HHV-6 and EBV demonstrate a significant specificity for RRMS (Almulla et al., 2024). For instance, the combination of heightened IgG and IgM responses targeting HHV-6 dUTPase resulted in a sensitivity of 87.3% and a specificity of 95.2% when comparing RRMS to healthy controls (Almulla et al., 2024). The significance of these findings lies in the fact that the dUTPases derived from both EBV and HHV-6 serve as biomarkers for assessing viral activity, replication, and the processes of reactivation or active “abortive” infection (Sommer et al., 1996; Tiwari et al., 2022). Furthermore, dUTPases function as pathogen-associated molecular pattern proteins (PAMPs), which may intensify immune pathology in a range of diseases, including MS (Parroche, 2011; Williams et al., 2016).

The prevalence of chronic fatigue, depression, and anxiety among patients with MS is notably high, with reported rates ranging from 75% to 90% for chronic fatigue, 27.01% to 50.0% for depression, and approximately 35.19% for anxiety (Ayache et al., 2022; Boeschoten et al., 2017; Feinstein et al., 2014; Nagaraj et al., 2013; Ormstad et al., 2020; Peres et al., 2022; Skokou et al., 2012). The onset of chronic fatigue syndrome and affective symptoms in RRMS may be elucidated, at least in part, by the interplay of autoimmune responses, activated immune-inflammatory pathways, and oxidative stress mechanisms (Almulla et al., 2023a; Koutsouraki et al., 2011; Maes et al., 2011; Mohr et al., 2001; Morris and Maes, 2013a; Morris et al., 2018; Ormstad et al., 2020). Furthermore, chronic fatigue and affective symptoms may detrimentally affect the patient’s quality of life and hinder the potential for remission, thereby influencing the overall prognosis of the condition.

Nonetheless, there exists a lack of data regarding the potential association between chronic fatigue syndrome and affective symptoms resulting from RRMS in relation to the replication or reactivation of EBV and/or HHV-6. Consequently, this investigation was conducted to explore the relationships between IgG, IgA, and IgM responses to EBNA, EBV-dUTPase, HHV-6-dUTPase, and the severity of chronic fatigue syndrome, depression, and anxiety in the context of RRMS.

## Subjects and Methods

### Subjects

A total of 58 patients diagnosed with RRMS were recruited for participation in this case-control study conducted at the Neuroscience Center of Alsader Medical City, located in Al-Najaf province, Iraq. The individuals were in the remitted phase of RRMS. The confirmation of the diagnosis of MS was achieved through the application of the McDonald criteria (Polman et al., 2011) and was conducted by a senior neurologist. Furthermore, a cohort of sixty-three healthy individuals from the same demographic region was recruited, encompassing hospital personnel, their acquaintances, and acquaintances of MS patients.

Patients and controls underwent a screening process designed to eliminate individuals who had experienced any lifetime diagnoses preceding the onset of MS of axis-I DSM-IV-TR neuropsychiatric disorders, including major depressive disorder (MDD), generalized anxiety disorder, obsessive-compulsive disorder, panic disorder, psycho-organic disorders other than MS, schizophrenia, post-traumatic stress disorder, substance use disorders (with the exception of nicotine dependence), bipolar disorder, and autism spectrum disorders. The study systematically excluded patients and controls who exhibited medical conditions, such as thyroid disorders, renal or liver diseases, diabetes mellitus type 1, cardiovascular diseases, (auto)immune disorders such as cancer, inflammatory bowel disease, rheumatoid arthritis, chronic obstructive pulmonary disease, psoriasis, and chronic fatigue syndrome/myalgic encephalomyelitis (CFS/ME). We also excluded controls and patients with neurodegenerative diseases encompassing conditions such as Parkinson’s disease and Alzheimer’s disease.

Written informed consent, duly documented, was secured from all patients diagnosed with RRMS, or their respective parents or legal guardians, in addition to control participants, prior to their enrollment in the study. The ethical approval for this research was granted by the institutional ethics board of the College of Medical Technology at The Islamic University of Najaf, Iraq, under document number 11/2021. Compliance with ethical standards was maintained through the observance of both Iraqi and international protocols, which encompass the World Medical Association Declaration of Helsinki, The Belmont Report, CIOMS guidelines, and the International Conference on Harmonization of Good Clinical Practice (ICH-GCP). The Institutional Review Board (IRB) adheres to the International Guidelines for Human Research Safety.

### Clinical Assessments

A senior neurologist undertook a semi-structured interview with the objective of collecting sociodemographic and clinical data. The neurologist employed the Expanded Disability Status Scale (EDSS) as delineated by Kurtzke (1983) to assess clinical disabilities (Kurtzke, 1983), alongside the Multiple Sclerosis Severity Score (MSSS) developed by Roxburgh, Seaman, and colleagues (2005) to evaluate the temporal progression of disability (Roxburgh et al., 2005).

Furthermore, we assessed the severity of the chronic fatigue, depression, and anxiety subdomains within the three months prior to the commencement of the study. Consequently, the Fibro-Fatigue scale (FF) (Zachrisson et al., 2002), the Hamilton Depression Rating Scale (HAMD) (Hamilton, 1960), and the Hamilton Anxiety Rating Scale (HAMA) (Hamilton, 1959) were employed to ascertain the severity of fibromyalgia-fatigue, depressive, and anxiety symptoms, respectively. We computed more specific symptom subdomain scores that reflect depressive and anxiety symptoms without physiosomatic symptoms, and physiosomatic symptoms without affective symptoms, based on our previous publications (Al-Hadrawi et al., 2023; Al-Hakeim et al., 2022; Almulla et al., 2021). The new term, “physiosomatic,” refers to psychosomatic symptoms while emphasizing that the symptom domain is linked to certain organic pathways (physio) rather than mental processes (psycho) (Maes et al., 2012). In order to achieve this objective, we calculated four distinct composite scores. a) Pure FF symptoms (pure FF) are the sum of the following FF items: “muscle pain,” “muscle tension,” “fatigue,” “autonomic symptoms,” “gastro-intestinal symptoms,” “headache,” and “flu-like malaise.” b) The sum of the HAMD items “depressed mood,” “feelings of guilt,” “suicidal ideation,” and “loss of interest” was used to calculate pure depressive symptoms (pure HAMD). c) The pure anxiety symptom domain (pure HAMA) was calculated as the sum of the HAMA items “anxious mood,” “tension,” “fears,” and “anxious behavior at interview.” d) A composite score based on z transformations was calculated for pure physiosomatic symptoms. This score was calculated by summing the z transformations of the HAMD items “anxiety somatic” + “gastrointestinal somatic” + “somatic general” + “genital” + the HAMA items “somatic muscular” + “somatic sensory” + “cardiovascular” + “respiratory symptoms” + “gastrointestinal” + “genitourinary” + “autonomic symptoms” + the FF symptoms “muscle pain” + “muscle tension” + “fatigue” + “autonomic” + “gastrointestinal” + “headache” + “a flu-like malaise.” The participant’s body mass index (BMI) was calculated by dividing their weight in kilograms by their height in meters squared.

### Biomarkers assays

Venipuncture with disposable instruments was employed to collect blood samples from fasting participants between 7:30 and 9:00 a.m. The blood was centrifuged at 3500 RPM for 10 minutes after being allowed to coagulate at room temperature for 15 minutes. The serum that was produced was aliquoted into Eppendorf containers for use in a variety of assays. The antibodies (IgA, IgG, IgM) specific to HHV-6-dUTPase, EBV-dUTPase, and EBNA-366-406 (EBNA) peptides, which were synthesized from Biosynthesis (Lewisville, TX, USA), were detected using an enzyme-linked immunosorbent assay (ELISA). The methodology for these procedures is extensively described in prior research (Almulla et al., 2023b; Trier et al., 2018). In summary, 100 microliters of various peptides were added to distinct wells of ELISA plates at a concentration of 5 micrograms per mL in 0.1 M carbonate buffer pH 9.5. Following incubation, rinsing, and blocking with 2% BSA, 100 microliters of the sera of the healthy controls and MS patients were added to duplicate wells at a dilution of 1:50 for IgA and 1:100 for IgG and IgM determination. Plates were subsequently rinsed, incubated, and secondary antibodies were subsequently added to each plate. The color development was measured, and indices were calculated using various sera as calibrators and controls after repetitive washing and the addition of substrate. As such, 9 biomarkers were employed in the analyses, namely IgG/IgA/IgM responses to EBNA, EBV- and HHV-6-dUTPases.

### Data analysis

Analysis of variance (ANOVA) was employed to compare continuous variables between study groups, whilst contingency table analysis was utilized to investigate the relationship between categorical variables. Point-biserial correlations were employed to examine the associations between scale and binary variables. Multiple comparisons and associations underwent p-correction for the False-Discovery Rate (FDR). The analysis employed Pearson’s product moment correlations to investigate the relationships among the scale variables. Multivariate regression analysis, with adjustments for demographic variables including age, gender, BMI, and smoking, was utilized to investigate the impact of immune reactivity to viral antigens on the scores of the neuropsychiatric rating scales (both total and pure rating scale scores). Both manual and stepwise methods were employed, with the latter utilizing entry and exit criteria established at p-values of 0.05 and 0.06, respectively. This analysis presented various model metrics, such as standardized beta coefficients, degrees of freedom (df), p-values, R², and F-statistics. In our investigation of heteroskedasticity, we employed the White test alongside the modified Breusch-Pagan test. Additionally, we assessed collinearity by analyzing tolerance and the variance inflation factor. All tests employed a two-tailed design, establishing a significance threshold of 0.05. In this study, we utilized IBM’s SPSS version 29 to conduct all statistical analyses.

Principal component analysis (PCA) was employed as a method for feature reduction, aimed at investigating the potential extraction of a singular “general” principal component from a collection of variables, such as the neuropsychiatric rating scores and the disability scores. This PCA must adhere to rigorous quality-fit standards, which encompass a Kaiser-Meyer-Olkin (KMO) value exceeding 0.6, the first principal component accounting for more than 50% of the variance, and all loadings on the first principal component being greater than 0.7. The principal statistical examination conducted was the multivariate analysis pertaining to neuropsychiatric rating scales in relation to the viral antigen markers. A preliminary power analysis conducted with G*Power 3.1.9.7 indicated that a minimum sample size of 73 individuals is required to investigate the R^2^ deviation from zero in a linear multiple regression (fixed model). This analysis is predicated on a power of 0.8, a significance level of 0.05, the inclusion of 4 covariates, and an effect size of 0.176.

## Results

### Demographic and clinical features of RRMS patients and controls

**Table 1** shows the demographic and clinical data of the RRMS patients and normal controls in this study. We found no differences observed in sex, age, BMI, education, and marital status between both study groups. There were somewhat more smokers among normal volunteers. All neuropsychiatric rating scale scores were significantly higher in RRMS patients than in controls.

**Table 1.** Sociodemographic and clinical data in patients with relapsing-remitting multiple sclerosis (RRMS) and healthy controls (HC).

| **Variables** | **Healthy Controls**<br>**(n=63)** | **RRMS Patients**<br>**(n=55)** | **F/$\chi^2$** | **df** | **p-value** |
| --- | --- | --- | --- | --- | --- |
| Age (years) | 31.4 (7.1) | 29.5 (6.3) | 2.18 | 1/116 | 0.142 |
| Sex (M/F) | 33/30 | 37/18 | 2.69 | - | 0.133 |
| BMI (kg/m<sup>2</sup>) | 26.33 (4.50) | 24.94 (3.52) | 3.40 | 1/116 | 0.068 |
| Education | 17.2 (5.1) | 18.4 (3.4) | 2.04 | 1/116 | 0.155 |
| Married/Separated | 26/37 | 31/24 | 2.67 | - | 0.139 |
| Smoking (N/Y) | 44/19 | 50/5 | 8.04 | - | 0.006 |
| DOI | - | 5.58 (4.94) | 80.31 | 1/116 | <0.0001 |
| MSSS | 0 | 1.71 (1.20) | MWU | - | <0.001 |
| EDSS | 0 | 1.03 (0.11) | MWU | - | <0.001 |
| Total FF score | 4.3 (3.5) | 23.6 (4.7) | 647.51 | 1/116 | <0.001 |
| Total HAMD score | 3.1 (2.2) | 11.7 (3.5) | 262 | 1/116 | <0.0001 |
| Total HAMA score | 4.4 (2.9) | 13.5 (3.6) | 228.93 | 1/116 | <0.0001 |
| PC FFAD (z scores) | -0.841 (0.371) | 0.964 (0.483) | 524.81 | 1/116 | <0.0001 |
| PC DISFADD (z scores) | -0.905 (0.129) | 1.03 (0.306) | 2102.10 | 1/116 | <0.0001 |
| Pure FF symptoms | 3.11 (2.86) | 17.81 (3.96) | 542.37 | 1/116 | <0.0001 |
| Pure HAMD symptoms | 0.825 (0.942) | 2.40 (1.14) | 66.879 | 1/116 | <0.0001 |
| Pure HAMA symptoms | 1.63 (1.46) | 3.90 (1.65) | 62.660 | 1/116 | <0.0001 |
| Pysiosomatic symptoms (z scores) | -8.24 (4.30) | 9.44 (4.82) | 443.401 | 1/116 | <0.0001 |

All results are shown as mean (SD). F: results of analysis of variance. $\chi^2$ : results of contingency analysis. MWU: Mann-Whitney U test.
DOI: duration of illness, MSSS: Multiple Sclerosis Severity Score; EDSS: Expanded Disability Status Scale; FF: Fibro-Fatigue scale; HAMD: Hamilton Depression Rating Scale; HAMA: Hamilton Anxiety Rating Scale; PC FFAD: first principal component extracted from FF, HAMD and HAMA scores; PC DISFADD: first PC extracted from EDSS, MSSS, FF, HAMD and HAMA scores.

### Construction of principal components

**Table 2** shows the outcome of different PCAs. First, we checked whether one PC could be extracted from the total FF, HAMD, and HAMA scores (see PCA #1). The first PC explained 90.01% of the variance and all loadings were higher than 0.934 (labeled PC FFAD from fibro-fatigue, anxiety, and depression). In PCA #2, we added the MSSS and EDSS scores. We were again able to extract one PC from these 5 scores which explained 84.88% of the variance, while all 5 variables showed high loadings (all > 0.870). This PC was labeled PC DISFFAD from disabilities, fibro-fatigue, anxiety, and depression. We were able to extract PCs, which complied with the a priori quality criteria, from the three IgG/IgA/IgM-EBNA-1 values (KMO=0.672, explained variance is 72.83%, labeled PC EBNA), three HHV-6 dUTPses (KMO=0.725, explained variance is 77.64%, labeled PC HHV-6-dUTPase), and three EBV dUTPases (KMO=0.675, explained variance is 73.34%, PC EBV-6-dUTPase). Consequently, we investigated whether one PC could be extracted from the three neuropsychiatric scores, EDSS, MSSS and the three viral PCs. Table 2, PCA #3 shows that one adequate PC could be extracted from these 8 variables with a good KMO value and explaining 75.58% of the variance. All eight variables loaded highly on this first PC and all loadings were higher than 0.800.

**Table 2:** Results of three different principal component analyses (PCA) conducted on disability scales and neuropsychiatric rating scales and immunoglobulin levels against viral antigens, including Epstein-Barr virus (EBV) and Human Herpesvirus-type 6 (HHV-6).

| **Features** | **PCA #1** | **PCA #2** | **PCA #3** |
| --- | --- | --- | --- |
| Total FF | **0.954** | **0.938** | **0.910** |
| Total HAMD | **0.934** | **0.916** | **0.842** |
| Total HAMA | **0.959** | **0.924** | **0.881** |
| MSSS | - | **0.870** | **0.877** |
| EDSS | - | **0.957** | **0.941** |
| PC EBNA | - | - | **0.800** |
| PC EBV-dUTPases | - | - | **0.861** |
| PC HHV-6-3dUTPases | - | - | **0.881** |
| KMO | 0.762 | 0.856 | 0.892 |
| % Variance | 90.007% | 84.883% | 76.576% |

FF: Fibro-Fatigue scale; HAMD: Hamilton Depression Rating Scale; HAMA: Hamilton Anxiety Rating Scale; MSSS: Multiple Sclerosis Severity Score; EDSS: Expanded Disability Status Scale; PC EBNA: first PC extracted from IgG/IgA/IgM responses to Epstein-Barr Virus nuclear antigen 1; PC EBV-dUTPases: first
PC extracted from IgG/IgA/IgM responses to EBV deoxyuridine-triphosphatase; PC HHV-6-3dUTPases: first principal component extracted from Human Herpes Virus type 6-dUTPase; KMO: Kaiser-Meyer-Olkin metric.

With the exception of IgG-dUTPase (r=-0.300, p=0.026, n=55, without false discovery rate p correction for multiple comparisons), no significant point-biserial correlations were observed between natalizumab administration and any viral data within the restricted cohort of RRMS patients. The administration of beta-interferon-1β was significantly correlated (without p-correction) with decreased levels of IgM-EBNA (r=-0.283, p=0.036), IgA-EBNA (r=-0.336, p=0.012), IgM-EBV-dUTPase (r=-0.305, p=0.023), and IgA-HHV-6-dUTPase (r=-0.267, p=0.049). However, all significances were nullified through the application of FDR p correction.

### Correlations between clinical ratings and immune responses to viral antigens

In all subjects combined, we found strong associations between PC DISFADD and the three PC scores (PC EBNA, PC EBV-dUTPases, and PC HHV-6-dUTPases), and the IgG/IgA/IgM responses to EBNA, EBV-dUTPases, and HHV-6-dUTPases (see **Table 3**). In the restricted RRMS patient sample, we found significant correlations between PC DISFFAD and the three PC scores, IgM/IgA-EBV-dUTPase, and IgA directed to HHV-6-dUTPase and EBNA.

**Table 3.**
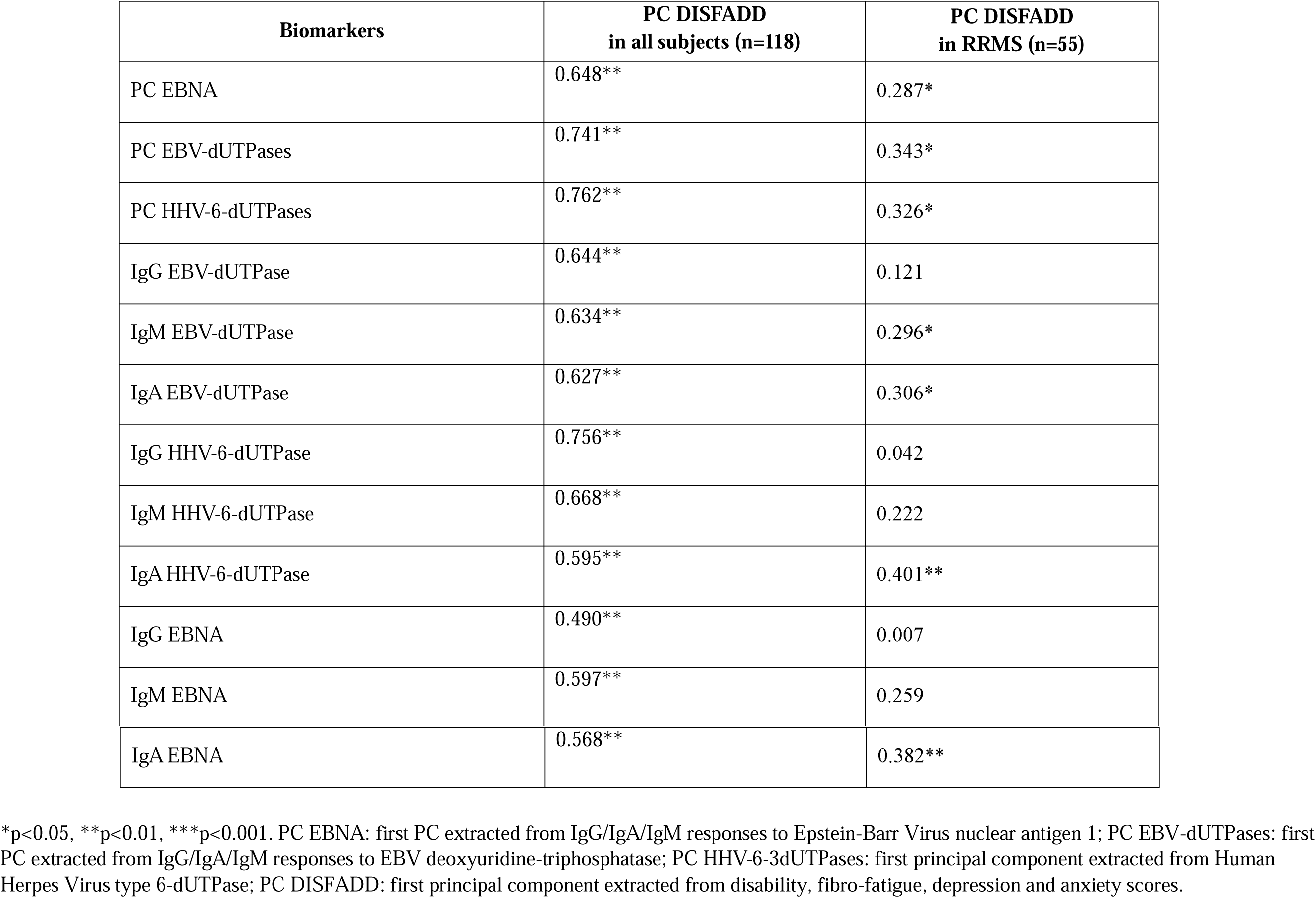
Intercorrelation matrix between between IgG, IgA and IgM against viral antigens and neuropsychiatric rating scales in relapsing-remitting multiple sclerosis (RRMS).

Consequently, we have also examined the associations between the three constructed “viral” PCs and the relevant rating scale scores (**Table 4**). We found significant correlations between the three viral PCs and the total FF, total HAMD, total HAMA scores, pure FF and physiosomatic scores, and pure HAMD and HAMA scores.

**Table 4.** Intercorrelation matrix between IgG, IgA and IgM against viral antigens and neuropsychiatric rating scales in relapsing-remitting multiple sclerosis.

| **Biomarkers** | **PC EBNA** | **PC EBV-dUTPases** | **PC HHV-6-dUTPases** |
| --- | --- | --- | --- |
| Total FF | 0.602<sup>**</sup> | 0.662<sup>**</sup> | 0.680<sup>**</sup> |
| Total HAMD | 0.502<sup>**</sup> | 0.592<sup>**</sup> | 0.589<sup>**</sup> |
| Total HAMA | 0.594<sup>**</sup> | 0.673<sup>**</sup> | 0.658<sup>**</sup> |
| Pure FF | 0.591<sup>**</sup> | 0.656<sup>**</sup> | 0.671<sup>**</sup> |
| Pure HAMD | 0.331<sup>**</sup> | 0.541<sup>**</sup> | 0.453<sup>**</sup> |
| Pure HAMA | 0.423<sup>**</sup> | 0.515<sup>**</sup> | 0.483<sup>**</sup> |
| Pure physiosomatic | 0.607<sup>**</sup> | 0.680<sup>**</sup> | 0.670<sup>**</sup> |

All $p < 0.001$ .
PC EBNA: first PC extracted from IgG/IgA/IgM responses to Epstein-Barr Virus nuclear antigen 1; PC EBV-dUTPases: first PC extracted from IgG/IgA/IgM responses to EBV deoxyuridine-triphosphatase; PC HHV-6-3dUTPases: first principal component extracted from Human Herpes Virus type 6-dUTPase; FF: Fibro-Fatigue scale; HAMD: Hamilton Depression Rating Scale; HAMA: Hamilton Anxiety Rating Scale.

### Immune responses to EBV and HHV-6 predict severity of neuropsychiatric symptoms

**Table 5** shows the results of multivariable regression analyses with the neuropsychiatric rating scale scores as dependent variables and IgG/IgA/IgM reactivity to viral antigens as explanatory variables while allowing for the effects of age, sex, BMI, and smoking. Nevertheless, these possible confounding variables were always non-significant. We found that IgG/IgM-HHV-6-dUTPAses explained together 63.7% of the variance in PC DISFADD in the total study group. IgG/IgM-HHV-6-dUTPAses significantly predicted the total scores of the three neuropsychiatric rating scales. IgG-EBV-dUTPase was additionally associated with the total HAMA score. A large part (38.7% -51.0%) of the variance in the pure FF, physiosomatic, HAMD and HAMA scores was predicted by a combination of the viral reactivation biomarkers. The pure FF score was best predicted by IgG/IgM-HHV-6-dUTPases, whereas the pure HAMD score was predicted by IgG/IgM-EBV-dUTPase. **Figure 1** shows the partial regression of the pure FF score on IgG directed against HHV 6-dUTPase. **Figure 2** shows the partial regression of the pure HAMD score on IgG-EBV-dUTPase. In the restricted study group of RRMS patients, we found that 16.1% of the variance in PC DISFFAD was explained by the regression on IgA-HHV-6-dUTPase (F=10.18, df=1/53, p=0.002, β=0.401). **Figure 3** shows the partial regression of PC DISFADD on IgA-HHV-6-dUTPase (after adjusting for sex and age).

**Figure 1.**
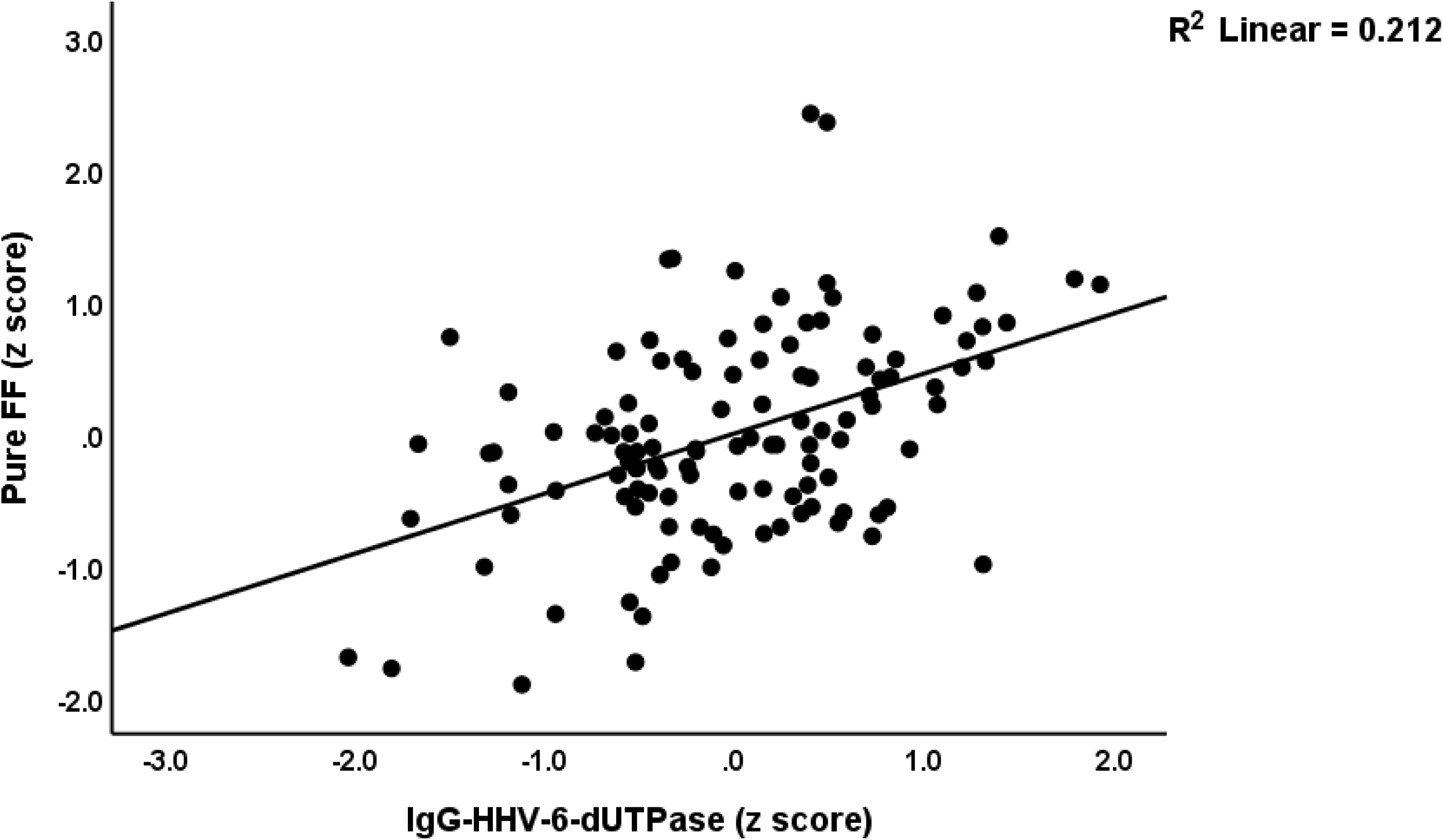
Partial regression of the pure physical subdomain of the Fibro-Fatigue (FF) scale on immunoglobulin (Ig)G directed against Human Herpesvirus-6 (HHVC6) deoxyuridine-triphosphatase (dUTPase); p < 0.001 (adjusted for age and IgM reactivity directed to HHV-6-dUTPase).

**Figure 2.**
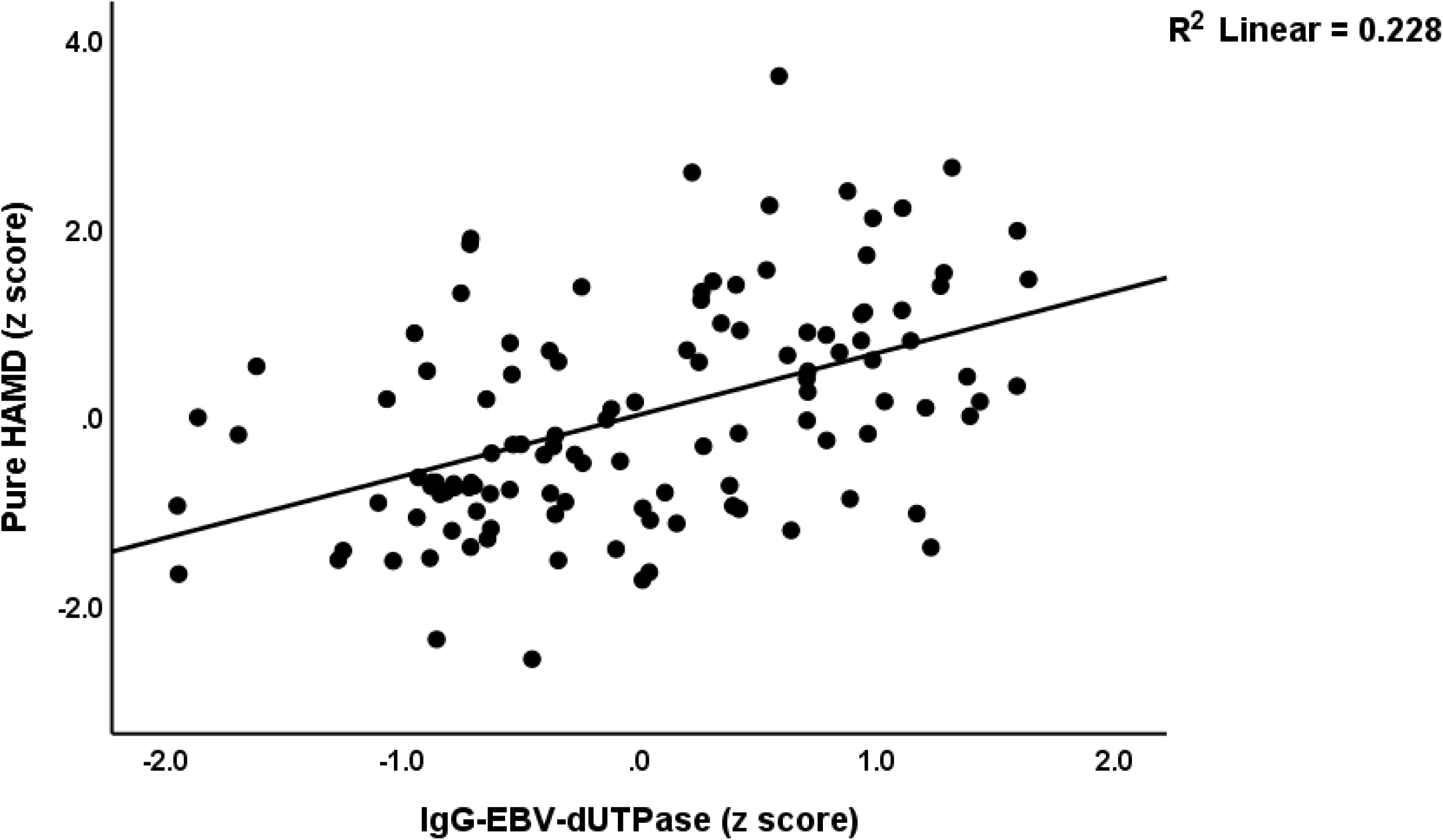
Partial regression of the cognitive subdomain of Hamilton depression rating scale on immunoglobulin (Ig)G directed against Epstein–Barr virus (EBV) deoxyuridine-triphosphatase (dUTPase); p<0.001 (adjusted for IgM-EBV-dUTPase).

**Figure 3.**
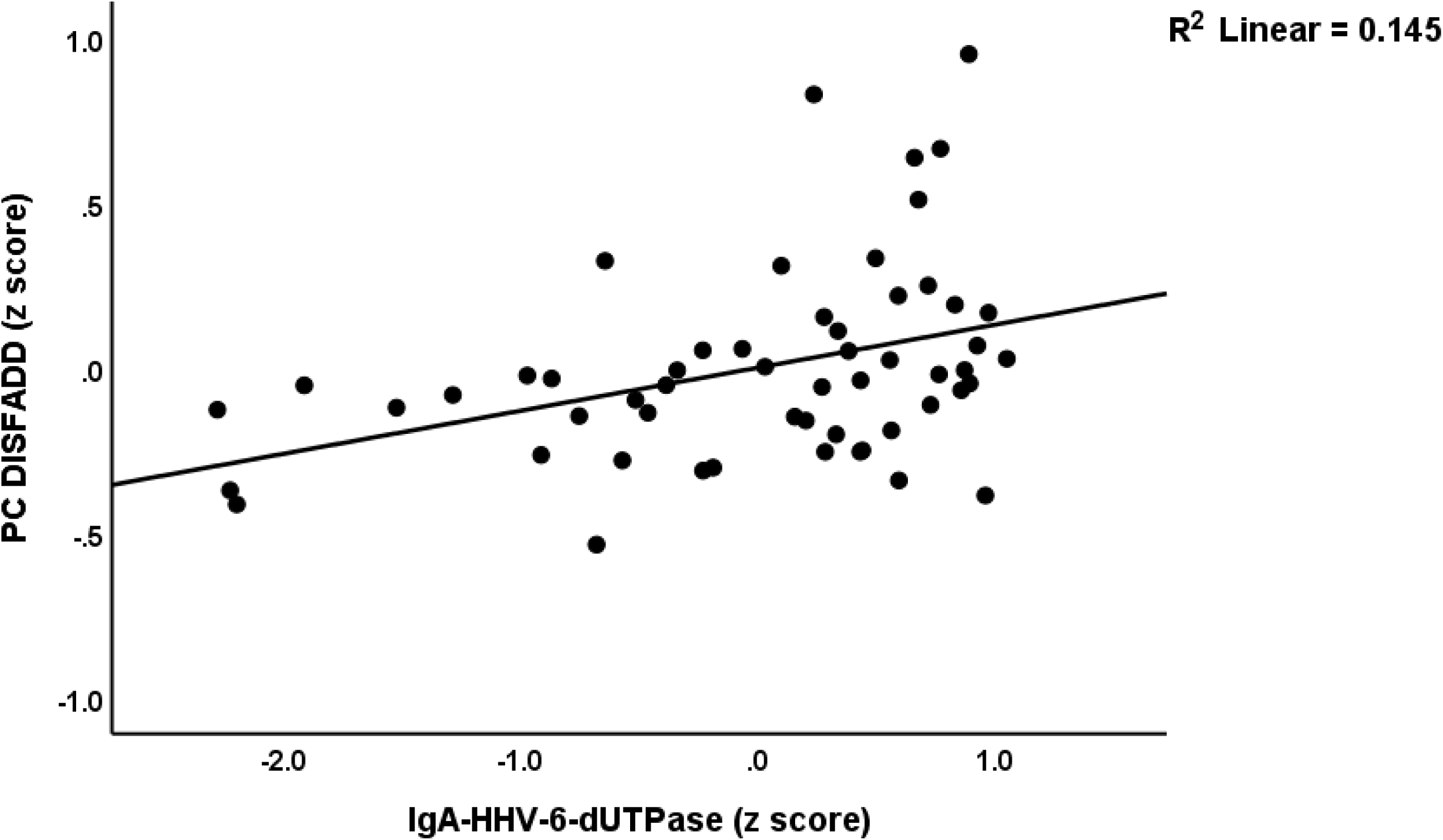
Partial regression of the first principal component extracted from disability, chronic fatigue, depression and anxiety rating scale scores (PC DISFFAD) on immunoglobulin (Ig)A directed against Human Herpesvirus-6 (HHV 6) deoxyuridine-triphosphatase (dUTPase); p=0.005 (adjusted for age). This regression is performed in RRMS patients only.

**Table 5.** Results of multiple regression analysis performed in the total study group with the affective symptoms profiles scores as dependent variables.

| Dependent Variables | Explanatory Variables | Coefficient statistics |  |  | Model statistics |  |  |  |
| --- | --- | --- | --- | --- | --- | --- | --- | --- |
| | | $\beta$ | t | p | R<sup>2</sup> | F | df | p |
| #1. PC DISFADD | **Model** |  |  |  | 0.637 | 100.80 | 2/115 | <0.001 |
|  | IgG-HHV-6-dUTPase | 0.554 | 7.76 | <0.001 |  |  |  |  |
|  | IgM-HHV-6-dUTPase | 0.326 | 4.56 | <0.001 |  |  |  |  |
| #2. Total FF | **Model** |  |  |  | 0.535 | 66.18 | 2/115 | <0.001 |
|  | IgG-HHV-6-dUTPase | 0.466 | 5.77 | <0.001 |  |  |  |  |
|  | IgM-HHV-6-dUTPase | 0.345 | 4.27 | <0.001 |  |  |  |  |
| #3. Total HAMD | **Model** |  |  |  | 0.402 | 38.67 | 2/115 | <0.001 |
|  | IgG-HHV-6-dUTPase | 0.479 | 5.23 | <0.001 |  |  |  |  |
|  | IgM-HHV-6-dUTPase | 0.214 | 2.34 | 0.021 |  |  |  |  |
| #4. Total HAMA | **Model** |  |  |  | 0.536 | 43.82 | 3/114 | <0.001 |
|  | IgG-HHV-6-dUTPase | 0.221 | 1.99 | 0.049 |  |  |  |  |
|  | IgM-HHV-6-dUTPase | 0.286 | 3.53 | 0.001 |  |  |  |  |
|  | IgG-EBV-dUTPase | 0.337 | 3.34 | 0.001 |  |  |  |  |
| #5. Pure FF | **Model** |  |  |  | 0.498 | 56.93 | 2/115 | <0.001 |
|  | IgG-HHV-6-dUTPase | 0.417 | 4.96 | <0.001 |  |  |  |  |
|  | IgM-HHV-6-dUTPase | 0.367 | 4.37 | <0.001 |  |  |  |  |
| #6. Pure HAMD | **Model** |  |  |  | 0.383 | 31.12 | 2/115 | <0.001 |
|  | IgG-EBV-dUTPase | 0.482 | 5.59 | <0.001 |  |  |  |  |
|  | IgM-EBV-dUTPase | 0.182 | 2.11 | 0.037 |  |  |  |  |
| #7. Pure HAMA | **Model** |  |  |  | 0.387 | 36.22 | 2/115 | <0.001 |
|  | IgG-EBV-dUTPase | 0.436 | 5.16 | <0.001 |  |  |  |  |
|  | IgM-HHV-6-dUTPase | 0.275 | 3.26 | 0.001 |  |  |  |  |
| #8. Pure physiosomatic | **Model** |  |  |  | 0.510 | 39.53 | 3/114 | <0.001 |
|  | IgG-HHV-6-dUTPase | 0.316 | 2.86 | 0.005 |  |  |  |  |
|  | IgM-EBV-dUTPase | 0.266 | 3.32 | 0.001 |  |  |  |  |
|  | IgG-EBV-dUTPase | 0.245 | 2.36 | 0.020 |  |  |  |  |

FF: Fibro-Fatigue scale; HAMD: Hamilton Depression Rating Scale; HAMA: Hamilton Anxiety Rating Scale; PC DISFADD: first principal component extracted from disability, FF, HAMD and HAMA scores; EBV-dUTPases: Epstein-Barr virus deoxyuridine-triphosphatase; HHV-6: Human Herpes Virus type 6-dUTPase.

## Discussion

### Increased chronic fatigue, depression, and anxiety symptoms in RRMS

The first finding of this study indicates that RRMS is marked by an exacerbation of chronic fatigue syndrome symptoms, alongside heightened symptoms of depression and anxiety. Nevertheless, the intensity of these three symptom domains is elevated only to a moderate extent.

Consequently, the average HAMD score among patients diagnosed with RRMS was recorded at 11.7 (SD=3.5). In contrast, outpatients exhibiting very mild MDD presented HAMD scores ranging from 15.2 (SD=6.3) to 17.7 (SD=6.5) (Vasupanrajit et al., 2024). In inpatients diagnosed with MDD, the average score on the HAMD is significantly elevated, specifically at 21.3 (SD=3.7) (Maes et al., 1986). In the current investigation, the average HAMA score was recorded at 13.5 (SD=3.6). In contrast, individuals diagnosed with MDD exhibit scores ranging from 27.2 (SD=2.6) to 29.5 (SD=1.8) (Maes et al., 1994). Furthermore, subjects experiencing minor depression present with a mean score of 15.6 (SD=1.4) (Maes et al., 1994). A similar pattern can be discerned in relation to the FF scale score. In our cohort of patients diagnosed with RRMS, the mean score on the FF scale was recorded at 23.6, with a standard deviation of 4.7. In individuals presenting with CFS/ME, the FF score is significantly higher, averaging approximately 43.3 (SD=7.7) (Maes et al., 2012). Nonetheless, the HAMD, HAMA, and FF scores obtained in patients with RRMS are either higher or comparable to the mean scores recorded in individuals experiencing a stable phase of schizophrenia, specifically ranging from 4.3 to 9.9, 5.3 to 16.3, and 5.8 to 18.2, respectively (Kanchanatawan et al., 2017). The significance of these affective and FF scores cannot be overstated, as they serve as the primary predictors of diminished quality of life among individuals diagnosed with schizophrenia (Kanchanatawan et al., 2019). It is noteworthy that the intensity of affective and chronic fatigue symptoms may escalate during acute relapses (Hanken et al., 2019; Khatibi et al., 2020; McCabe, 2005). However, it is also important to consider that some authors have documented non-significant associations between affective symptoms and the various clinical phases (Jefferies and Lambon Ralph, 2006).

It is crucial to note that the current study computed sub scores that evaluated pure cognitive depressive and anxiety symptoms in isolation from chronic fatigue and physiosomatic symptoms. Furthermore, given that we evaluated the HAMD, HAMA, and FF scales, as well as their cognitive and physiosomatic subdomains, within the three months preceding the study, it is reasonable to infer that the remitted phase of RRMS is associated with chronically elevated ratings of chronic fatigue syndrome, physiosomatic and affective symptoms. Given that patients with lifetime diagnoses of MDD, as well as CFS/ME, were excluded, it is reasonable to infer that RRMS is defined by new onset chronic fatigue syndrome and affective symptoms.

The current study reveals a significant finding: a singular latent construct can be derived from the affective and chronic fatigue symptoms, as well as the EDSS and MSSS scores. This suggests that the interconnected relationships between heightened motor and sensory impairments, along with increased chronic fatigue and affective symptoms, represent highly interrelated manifestations of RRMS. This further emphasizes that these symptoms and disabilities exhibit shared underlying pathways.

### Neuropsychiatric scores are predicted by viral replication

The second significant discovery of this study is the significant correlation between PC DISFFAD, the severity of depression, anxiety, and chronic fatigue symptoms, and the markers of HHV-6 and EBV reactivation, replication, or active “abortive” infection (Sommer et al., 1996; Tiwari et al., 2022) we measured in our study. In addition, we were able to extract a single latent construct from the viral reactivation and replication biomarkers, as well as the disability, affective, and chronic fatigue scores. This not only demonstrates a strong correlation between the viral biomarkers, disabilities, and neuropsychiatric scores in RRMS, but also indicates that HHV-6 and EBV reactivation/replication are critical components of this disorder.

Prior research indicates that FFAD symptoms during acute relapses may be ascribed to activated immune-inflammatory and oxidative stress pathways (Moore et al., 2012; Morris and Maes, 2013a; Ormstad et al., 2020; Šabanagić-Hajrić et al., 2015). For example, elevated levels of IL-6 are positively correlated with the depressive symptoms associated with RRMS (Kallaur et al., 2016; Koutsouraki et al., 2011). Kallaur et al. (2016) demonstrated that depression is a manifestation of the neurological impairments associated with multiple sclerosis and that both symptom categories are predicted by indicators of peripheral immunological activation (Kallaur et al., 2016). In the relapsing phase of RRMS, FFAD symptoms are significantly correlated with several inflammatory and anti-inflammatory cytokines, particularly those of the T helper (Th)17 axis (Almulla et al., 2023a). Consequently, it was determined that the activation of immune-inflammatory, autoimmune, and oxidative pathways, along with the resultant damage to CNS structures in MS, may heighten susceptibility to affective symptoms and chronic fatigue syndrome (de Carvalho Jennings Pereira et al., 2020; Feinstein, 2004; Morris and Maes, 2013a). It is essential to emphasize that affective disorders, CFS/ME are marked by activated immune-inflammatory and oxidative stress pathways (Maes, 2023; Maes and Carvalho, 2018; Morris and Maes, 2013b).

Consequently, the hypothesis posits that heightened neurotoxicity resulting from immune activation (e.g., elevated M1 macrophages, Th1 and Th17 cytokines) in RRMS may impair affective circuits and energy homeostasis in the CNS, consequently leading to affective symptoms, chronic fatigue, and psychosomatic manifestations (Maes and Carvalho, 2018; Morris and Maes, 2013a). It is crucial to note that dUTPases may function as PAMPs and activate Toll-Like receptors, nuclear factor-κB, and M1 and Th-1 cytokine production, thereby exacerbating the pathogenesis of MS (Ariza et al., 2022; Parroche, 2011; Waldman et al., 2008; Williams et al., 2016; Williams et al., 2023; Zhang et al., 2017). HHV-6 seropositivity and reactivation are linked to altered levels of pro-inflammatory cytokines, including Th1 cytokines (Chapenko et al., 2003). Additionally, autoimmune responses and the resulting tissue injury observed in RRMS may be exacerbated by the reactivation and replication of EBV and HHV-6, particularly through cross-reactivity or mimicry (Fotheringham and Jacobson, 2005; Lanz et al., 2022). Consequently, HHV-6 and EBV reactivation may not only sustain ongoing immune activation during the remission phase of RRMS (Almulla et al., 2024) but also appear to drive the affective symptoms and chronic fatigue syndrome associated with RRMS.

## Limitations

The findings of the present investigation warrant replication in other nations and cultures. The results could have been more intriguing if we had also evaluated the viral biomarkers and neuropsychiatric rating instruments during acute relapses. In order to evaluate biomarkers and rating scales during the acute relapse phase and remission, future research should employ a prospective design. Although some people may consider that the study was performed using a smaller study sample, the sample size was estimated a priori based on a power of 0.8. Moreover, the power of 1.0 was obtained through the post-hoc computation of the achieved power in the primary statistical analysis, which involved multiple regression with PC DISFADD as the dependent variable and the viral biomarkers as explanatory variables.

## Conclusions

This study demonstrates a significant association between heightened IgG/IgA/IgM reactivity to EBNA, EBV-dUTPase, and HHV-6 dUTPase with PC DISFFAD, as well as the severity of depression, anxiety, and chronic fatigue syndrome in the context of RRMS. A significant portion of depression and anxiety symptoms, which are distinct from psychosomatic symptoms (38.3%-38.7%), as well as psychosomatic symptoms themselves (51 %), is anticipated to be influenced by immune reactivity to HHV-6-dUTPase and EBV-dUTPase. The findings suggest that the reactivation, replication, or active “abortive” infection of HHV-6 and EBV plays a significant role in the initiation or persistence of disabilities, affective symptoms, and chronic fatigue syndrome associated with RRMS.

## Data Availability

The corresponding author (MM) is prepared to grant access to the file associated with this study upon receipt of a valid request and subsequent to a comprehensive analysis of the data set.

## Acknowledgements

The authors are deeply grateful to the Neuroscience Center of Alsader Medical City in Al-Najaf province, Iraq, for their invaluable support in the data collection process.

## Ethical approval and consent to participate

The Ethics Committee of the College of Medical Technology at the Islamic University of Najaf, Iraq, granted sanction for the investigation (Document No. 11/2021). Written informed consent was obtained from all patients and control participants, and all procedures were conducted in accordance with Iraqi and international ethical standards.

## Author’s contributions

AFA oversaw the blood sample collection and patient-related procedures. AV and AFA conducted the serum biomarker quantification. The statistical analysis was conducted by MM. Visualization was executed by MM. MM authored the initial manuscript, which was subsequently revised by AFA, EV, ED, DS, YZ, and AV. All authors authorized the most recent version.

